# Estimating the local spatio-temporal distribution of disease from routine health information systems: the case of malaria in rural Madagascar

**DOI:** 10.1101/2020.08.17.20151282

**Authors:** Elizabeth Hyde, Matthew H. Bonds, Felana A. Ihantamalala, Ann C. Miller, Laura F. Cordier, Benedicte Razafinjato, Herinjaka Andriambolamanana, Marius Randriamanambintsoa, Michele Barry, Jean-Claude Andrianirinarison, Mauricette A. Nambinisoa, Andres Garchitorena

## Abstract

**Background:** Reliable surveillance systems are essential for identifying disease outbreaks and allocating resources to ensure universal access to diagnostics and treatment for endemic diseases. Yet, most countries with high disease burdens rely entirely on facility-based passive surveillance systems, which miss the vast majority of cases in rural settings with low access to health care. This is especially true for malaria, for which the World Health Organization estimates that routine surveillance detects only 14% of global cases. The goal of this study was to estimate the unobserved burden of malaria missed by routine passive surveillance in a rural district of Madagascar to produce realistic incidence estimates across space and time, less sensitive to heterogeneous health care access.

**Methods:** We use a geographically explicit dataset of the 73,022 malaria cases confirmed at health centers in the Ifanadiana District in Madagascar from 2014 to 2017. Malaria incidence was adjusted to account for underreporting due to stock-outs of rapid diagnostic tests and variable access to healthcare. A benchmark multiplier was combined with a health care utilization index obtained from statistical models of non-malaria patients. Variations to the multiplier and several strategies for pooling neighboring communities together were explored to allow for fine-tuning of the final estimates. Separate analyses were carried out for individuals of all ages and for children under five. Cross-validation criteria were developed based on overall incidence, trends in financial and geographical access to health care, and consistency with geographic distribution in a district-representative cohort. The most plausible sets of estimates were then identified based on these criteria.

**Results:** Passive surveillance was estimated to have missed about 4 in every 5 malaria cases among all individuals and 2 out of every 3 cases among children under five. Adjusted malaria estimates were less biased by differences in populations’ financial and geographic access to care. Average adjusted monthly malaria incidence was nearly four times higher during the high transmission season than during the low transmission season. Geographic distribution in the adjusted dataset revealed high transmission clusters in low elevation areas in the northeast and southeast of the district that were stable across seasons and transmission years.

**Conclusions:** Understanding local disease dynamics from routine passive surveillance data can be a key step towards achieving universal access to diagnostics and treatment. Methods presented here could be scaled-up thanks to the increasing availability of e-health disease surveillance platforms for malaria and other diseases across the developing world.

## BACKGROUND

The lack of big data analytics in global health care delivery represents an enormous gap preventing progress toward universal health coverage^1^. The realm of infectious diseases is a prime target for the application of these methods, as increasingly available spatial and temporal information can be harnessed in combination with epidemiological models to produce precise estimates of disease burdens^2,3^. The most common data sources used to understand burdens of endemic diseases are routine facility-based health management information systems (HMIS) and household surveys. HMIS data have some degree of clinical and temporal granularity and are useful for health planning, but do not provide accurate information on disease burdens because they are only representative of those who access health care. In comparison, nationally representative household surveys (e.g. Demographic and Health Surveys) are heavily relied on for tracking development targets and establishing control priorities, but their data are clinically and spatio-temporally coarse (they are collected every 5 years, in samples that are representative of large regions), and involve limited diagnostic tests. Designated surveillance sites can add high quality data in particular locations, but are expensive and not scalable for localized planning. The prevailing approach for bridging this space is in the form of precision health mapping, where health outputs from coarse epidemiological data are fit from much more granular geospatial environmental data^4-6^. Though this approach produces projections at fine spatio-temporal scales over large geographic areas, these cannot be used by district managers for local planning due to limited accuracy. This represents a significant missed opportunity, because health systems are sitting on enormous quantities of granular data that could be used for local disease control if systematic biases in these data could be addressed.

Malaria is a good example of the challenges and opportunities in the use of health system data for disease control. Despite being preventable and treatable, malaria continues to cause an estimated 228 million cases and 405,000 deaths worldwide each year ^7^. Widespread implementation of malaria control measures such as insecticide-treated bed net distribution and indoor residual spraying has resulted in a steady decrease of global incidence, but this trend has recently slowed and even reversed in some areas ^,9^. Universal access to rapid diagnosis and treatment is a key strategy to reduce the burden of malaria, but access to health care remains stubbornly low in rural areas of Sub-Saharan Africa where most of the burden accumulates^9^. In 2017, only one third of African children with fever were brought to a medical provider. Thus, a substantial number of malaria cases were not diagnosed, treated, or included in surveillance statistics^9^. This could be worsened under the current COVID-19 pandemic, which is disrupting supply chains, community health and outreach activities, and could further undermine access to health facilities due to the stigma associated with COVID-19^10,11^.

Surveillance is critical for both disease control and elimination, and has become one of the three pillars of the Global Technical Strategy for Malaria 2016-2030^12^. Most malaria control programs rely on passive surveillance systems via case detection at health facilities. Yet, passive surveillance is known to grossly underestimate the incidence of malaria^13-16^ because only symptomatic patients who seek care at health facilities are recorded. In 2012, the World Health Organization estimated that only 14% of malaria cases worldwide were detected with routine surveillance^17^. Even in countries committed to malaria elimination, nearly two thirds of cases are missed by national surveillance systems^18^. Passive surveillance is especially unsuited to estimate local malaria burdens for remote populations in rural areas, as health centers are sparsely distributed and health care utilization tends to decrease exponentially as distance to a health facility increases ^19-22^. Active surveillance can enhance case detection, but its application remains limited to near-elimination areas due to resource constraints^23^. Thus, innovations are needed to improve the use of passive surveillance data in high transmission areas in order to increase the ability of local control programs to track disease dynamics within a health district, efficiently deploy resources, and target interventions to high-risk populations.

Implementation of such methods could be particularly important in Madagascar, one of the poorest countries in the world with one of the least-funded health systems^24,25^. Malaria remains one of the leading causes of mortality in the island^26^, with 22.4 of its 25.6 million people living in areas with high transmission^27^. Madagascar is one of only seven countries in the world where malaria incidence and mortality rates increased by more than 20% in 2015 compared to 2010 levels. Between 2016 and 2017, the country saw an increase of more than half a million cases^8^. Yet, during that time, only 15.5% of children with reported fever had an RDT done and only 10.1% were treated with an antimalarial^28^. Access to healthcare is particularly low in rural areas of the country, where over three quarters of the population live^29^. In 2014, the Ministry of Health (MoH) partnered with the healthcare NGO PIVOT to strengthen the rural health district of Ifanadiana, located in southeastern Madagascar where malaria transmission is highest^30^, to serve as a model health system for the country. In support of local malaria control efforts, the goal of this study was to estimate the burden of malaria missed by routine passive surveillance to help produce more realistic estimates of malaria incidence across space and time, less sensitive to changes in health care access. For this, we used a geographically-explicit patient dataset from the district’s health centers and we adjusted malaria estimates following a detailed characterization of health care utilization drivers in non-malaria patients.

## METHODS

### Study site

Ifanadiana is a rural district located in the Vatovavy-Fitovinany region in Madagascar. According to the MoH, Ifanadiana contained approximately 195,000 people in 2015, the vast majority of whom subsist on agriculture (84.8%)^29,31,32^. The district is divided into 13 communes (subdivisions with approximately 15,000 people each), which are further divided into 195 Fokontany (the smallest administrative unit, containing one or several villages). Ifanadiana has one reference hospital, one major public health center (CSB2) in each of its 13 communes, and six additional basic health centers (CSB1) in the larger communes (Figure 1). Passive malaria surveillance is continuously conducted at all of the 19 public health centers throughout the Ifanadiana District, aggregated from routine health registries of clinical patients.

**Figure 1.**
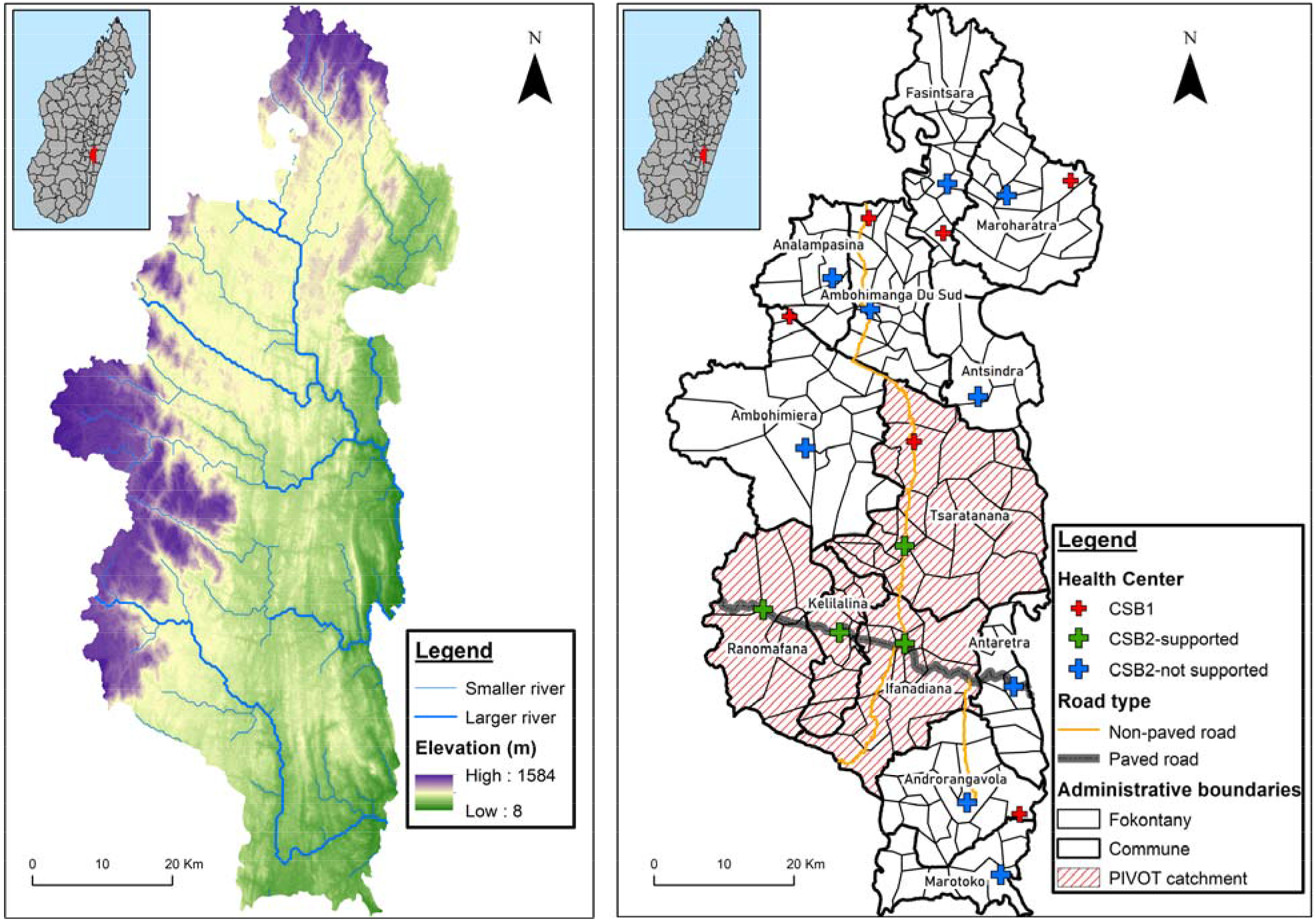
Map of the Ifanadiana district in Madagascar. The left panel shows elevation and waterways. The right panel shows administrative boundaries, roads, health centers (CSBs), and the PIVOT initial catchment area.

In 2014, a baseline study indicated that Ifanadiana had some of the highest poverty rates and worst health indicators in Madagascar. Nearly three fourths of the population lived in extreme poverty. The mortality rate for children under five was 145 deaths per 1000 live births, more than double the national estimate of 62 per 1000^31,33^. Malaria prevalence in the area where the district is located is the highest in the country, with prevalence ranging from 6 to 18%^30^. While more than a third of children under five in Ifanadiana had reported fever in the previous two weeks, only 42% were taken to a health center^34^. Low access to health care was strongly associated with substantial financial and geographic barriers^35^. For instance, only one fourth of the population lives within an hour’s travel of a health center^36,37^.

Since 2014, PIVOT has supported the public health system of Ifanadiana at all levels (hospital, health centers and community health workers) guided by the WHO framework for health system strengthening^38^. The intervention initially covered approximately one third of the district’s population. In these areas, PIVOT has helped remove financial barriers to care; improved readiness at health facilities, which includes personnel (quantity of staff and training), supply chain (equipment and consumable), infrastructure, and health management information systems; created an ambulance network; and implemented clinical programs that target tuberculosis, malnutrition and childhood illness through strengthened programs at all levels of care. Following PIVOT’s support, the number of cases of malaria diagnosed at health centers in these areas experienced a sudden increase due to rapid improvements in overall health care utilization^35,39^. To further support local malaria control programs, PIVOT aims to support the MoH to optimize interventions geographically in a context of heterogeneous disease burdens.

### Data collection

Data was obtained from health center registers on all individuals who visited a public health center for an outpatient consultation in Ifanadiana district between January 2014 and December 2017. Every 3-4 months, each public health center was visited and digital photos were taken of the register, in agreement with the head of the public health center. A digital database of register photos was created and stored in a secure server. De-identified information including age, Fokontany of residence, and malaria status of each new patient was entered into an electronic database (follow-up visits were excluded). Health center staff made malaria diagnoses with rapid diagnostic tests (RDTs) for patients presenting with fever, following national guidelines.

In addition to health system information, data from the I-HOPE cohort was used to estimate the geographic distribution of fever prevalence by age group in Ifanadiana^34^. The I-HOPE longitudinal cohort study, representative of the population in Ifanadiana district, was initiated in 2014 to understand the evolution of health and socio-economic characteristics as one of the information pillars to create a model health district. It consists of a series of biannual surveys conducted by INSTAT on the same households over time, with questionnaires and methods adapted from the Demographic and Health Surveys and other international surveys. The survey has a two-stage stratified sampling design covering 1, 600 households (~8000 people) in 80 geographic clusters across the district. Information from the cohort, which was available for 2014 (April-May), 2016 (August-September) and 2018 (April-May), included questions to assess reported fever among children under five years (previous two weeks) and among all household members (previous four weeks).

To obtain per capita estimates, population data for each Fokontany were obtained from the MoH. The population of children under five years old was estimated at 18% of the total population, per the MoH. Data on monthly stocks of RDTs at the end of each month and number of days with RDT stock-outs were obtained from each health center’s monthly report to the district. Use of MoH data for this study was authorized by the Secretary General of the MoH, the Medical Inspector of Ifanadiana district, and Harvard Medical School IRB. The I-HOPE cohort study was approved by the Madagascar National Ethics Committee and Harvard Medical School IRB.

Finally, we used a geographic information system containing data on locations of all health centers, more than 20,000 km of footpaths, over 100,000 buildings, and nearly 5,000 residential areas in the district. This was obtained following a participatory complete mapping of Ifanadiana in 2018-2019, from very high resolution satellite images available through OpenStreetMap^37^. This data was queried on QGIS via the QuickOSM plugin and was used to estimate shortest path distances between health centers and each Fokontany.

### Data analysis

Patient-level information from each health center was aggregated to estimate per capita utilization rates and malaria incidence per month for each Fokontany in Ifanadiana district. In order to obtain more realistic estimates of malaria incidence per Fokontany-month, malaria incidence was adjusted to account for underreporting due to stock-outs of RDTs and variable access to healthcare due to geographic and financial barriers. Access-based adjustments were based on a benchmark multiplier method that used a healthcare utilization index, which was obtained from a model of Fokontany per capita utilization in non-malaria patients. Several multipliers and pooling strategies were explored to improve adjustments in the Fokontany with the lowest access to care, and the most plausible set of estimates was identified by comparing each set of estimates according to four evaluation criteria (next section). Each of these steps is explained in detail below.

To account for the reduction in malaria diagnoses as a result of health center stock-outs of RDTs, each Fokontany was matched with its nearest health center. The shortest path distance between all health centers and Fokontany (average distance for every house) was estimated via the Open Source Routing Machine (OSRM) engine. The number of diagnosed malaria cases in health centers and months with stock-outs (14 months) were adjusted by a factor inversely proportional to the number of days in the month when RDTs were available. For example, if RDTs were only available at a health center for half of the month, the adjusted malaria cases for that month would be double the original reported number. In months in which stock-outs persisted for an entire month at a given health center (10 months), we assigned missing values for the malaria incidence in all Fokontany served by that health center.

To account for the effect of low health care access on malaria incidence, we used results from a spatio-temporal model of health care utilization in Ifanadiana during the same study period. Details on this model are published elsewhere^36^, and model specifications are available in the Appendix. Briefly, per capita health center utilization rates for each Fokontany were modeled using Binomial regressions in generalized linear mixed models, with a random intercept introduced for the closest health center. The model accounts for the non-linear effect of travel distance from each Fokontany to the nearest health center; the impact of programs implemented to reduce financial and geographic barriers; linear and seasonal trends in utilization rates in the absence of those programs; baseline differences in the type of health center (CSB1 or CSB2); and the number of health staff over time in the closest health center^36^. Based on model predictions for non-malaria patients, a health center utilization index was produced for each Fokontany-month in Ifanadiana, scaled between zero (no access; set at zero consultations per person-month) and one (full access; set at 0.166 consultations per person-month, equivalent to 2 consultations per person-year, excluding malaria).

A simplified benchmark multiplier method was used to adjust malaria incidence with the health care utilization index produced from non-malaria patients. This method combines information about the known members of a target population (the benchmark; for example, the number of people with malaria who are diagnosed at a health center) with the proportion of the target population that appears in the benchmark (for example, the proportion of people with malaria who go to a health center)^40^. The reciprocal of the proportion is called the multiplier. The true size of the target population (in this case, the true number of people with malaria in Ifanadiana) is estimated as the product of the benchmark and the multiplier. Populations with the best health care access (i.e. located very close to a health center with fee-exemptions in place) are not adjusted, while populations with the worst access are adjusted using the largest multiplier (Figure 2). The simplified benchmark multiplier formula is defined as:

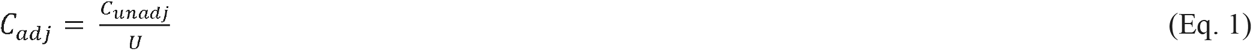

**Figure 2.**
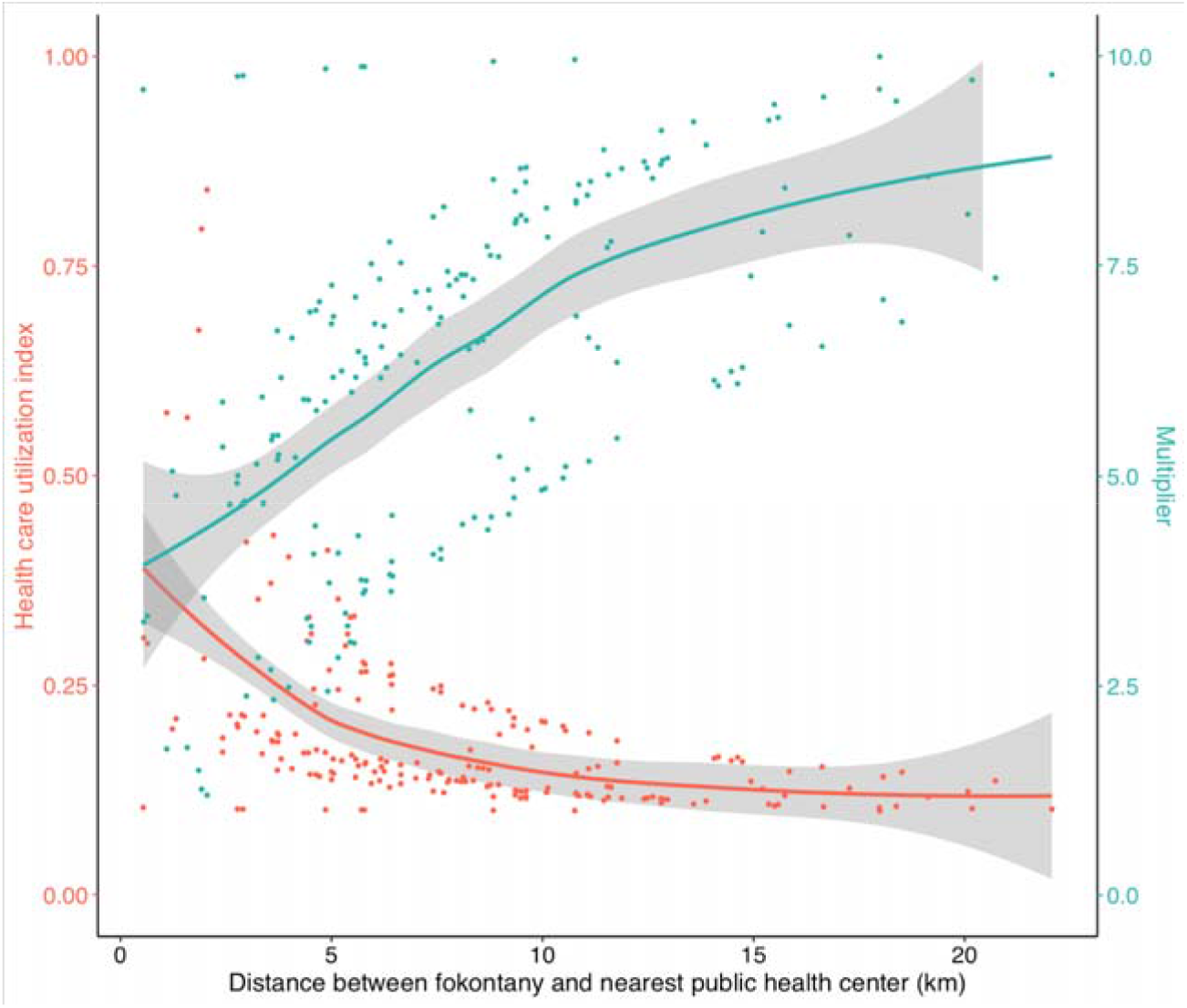
Illustration of benchmark multiplier adjustments to passive malaria surveillance data using a health care utilization index. Each dot represents the average health care utilization index (orange) or resulting multiplier (teal) for one of the 195 Fokontany in Ifanadiana over the study period. In this example, average per capita health care utilization index is normalized from 0.1 to 1, where the maximum is equivalent to 2 visits per year (excluding malaria). Both variables are plotted as a function of distance between each Fokontany and its nearest health center. The solid lines are smoothed conditional means (LOESS method) and the grey areas are the 95% confidence intervals. Fokontany with smaller health care utilization indices have larger multipliers, resulting in greater adjustments after the benchmark multiplier method was applied.

Where C_unadj_ represents the unadjusted monthly cases in a given fokontany for a given month, U represents the health care utilization index for the fokontany from the model described above, and C_adj_ represents the resulting adjusted monthly cases in the fokontany for the month. Multiplying the benchmark by the multiplier (1/U) can result in drastic changes in magnitude of the resulting estimates. To account for this, the lower limit of the health care utilization index was varied from 0.01 to 0.2 in steps of 0.01, with the upper limit remaining one. This allowed for fine-tuning of the adjusted monthly malaria incidence estimates.

Finally, due to extremely low access to care, several Fokontany had no malaria cases for several months even during the high transmission season, particularly those located at farther distances (e.g. 10-20 km) from health centers. For instance, 37 of the 195 Fokontany did not have any malaria cases during more than half of the high season months (December to May) in the four years of the study, 86% of which were further than 5 kilometers from a health center. Because Fokontany that have a malaria incidence of zero during a given month cannot be adjusted using a multiplier, we explored several strategies to pool the number of malaria cases in these Fokontany with the cases in neighboring Fokontany and estimate a pooled incidence that could then be adjusted for low health care access. We explored pooling with the *k*-nearest neighbors (2, 3, 4 and 5) and with neighbors within a certain distance (3, 4, and 5km).

The combination of 8 different pooling strategies and 21 different lower limits set for the health utilization index resulted in 168 alternative sets of adjusted malaria incidence estimates, both for individuals of all ages and for children under five.

### Evaluation of model estimates

The lack of a district-representative active surveillance survey during the study period meant that alternative sets of adjusted estimates of malaria incidence from passive surveillance could not be robustly compared to an unbiased training dataset for validation. We established four evaluation criteria to choose the most plausible set of incidence estimates in Ifanadiana based on the available data (Table 1). This was done both for individuals of all ages and for children under 5.

**Table 1.**
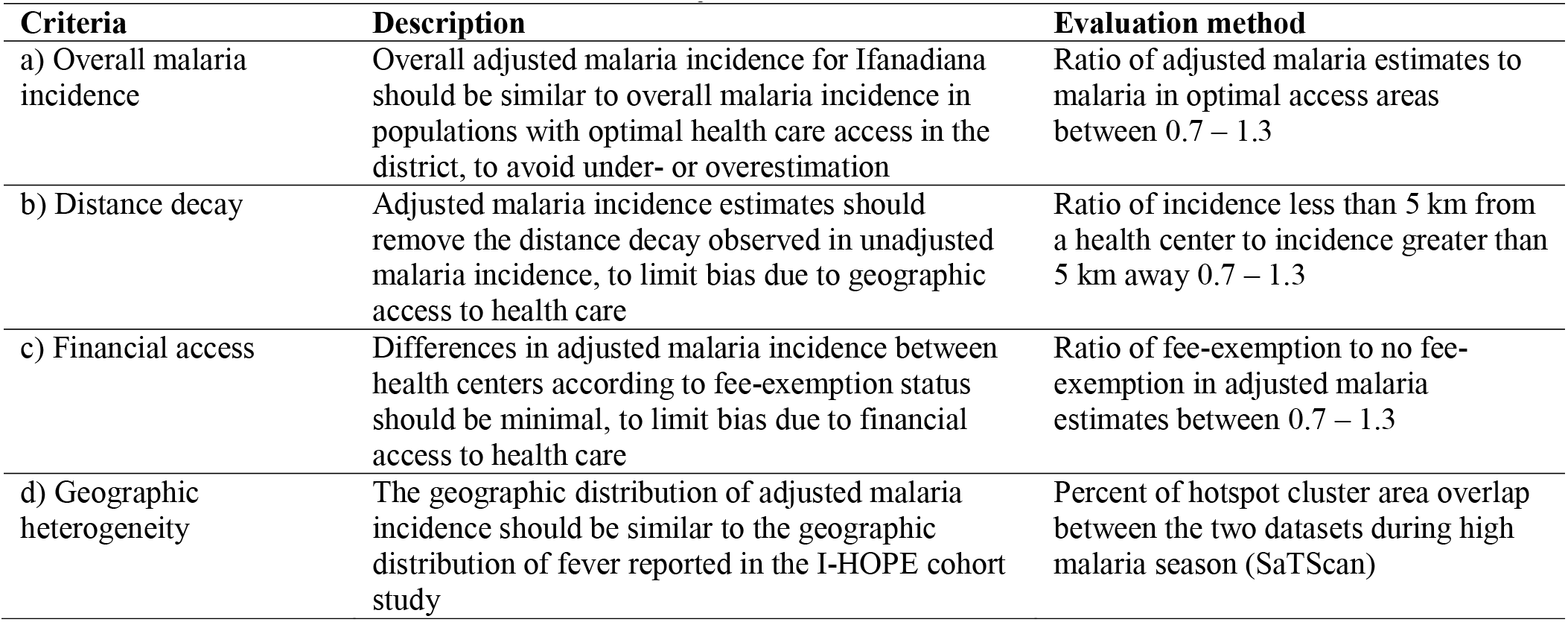
Evaluation criteria for alternative sets of adjusted malaria incidence estimates.

Evaluation criteria are based on: a) consistency of *overall malaria incidence* in the district with incidence in areas with optimal access to healthcare; b) reduction of *distance decay* relationship; c) reduction of bias due to *financial access* to care; and d) consistency of *geographic heterogeneities* in district with patterns observed in the I-HOPE cohort study. The first three criteria rely on the assumption that the burden of malaria in populations with good access to health care (e.g. those who live near health centers, or in areas where user fees have been removed) is similar to the burden elsewhere because the per capita distribution of malaria is predominantly driven by ecological and epidemiological factors, and not by health care access^41-44^. Although health centers diagnose and treat malaria patients, the main malaria prevention activities in Madagascar (e.g. bed net distribution, indoor residual spraying) that could affect transmission are delivered through mass-campaigns to all at-risk populations.

#### Overall malaria incidence

To avoid under- or overestimation of overall malaria incidence in the district, we assumed that adjusted estimates should be similar to unadjusted malaria incidence in populations with optimal access to health care. These were defined as populations from Fokontany that are in close proximity (≤ 2.5 km) to a PIVOT-supported health center following initial implementation of health system strengthening interventions. These populations travelled short distances to care and benefited from improved facilities, with greater staffing, and point-of-care fees for most health services removed. They represented a total population of 10,583 individuals of all ages, including 1,905 children distributed across 4 Fokontany in 4communes, with an average health system utilization index of 0.66 (on a scale from 0 to 1). The 4-year annual malaria incidence average in this population for 2014-2017 was 397 cases per 1000 population among individuals (33 cases per 1000 population per month) and 631 cases per 1000 population among children under five (53 cases per 1000 per month). To assess this criterion, we estimated the ratio of average malaria incidence in each adjusted dataset to average malaria incidence in the higher access dataset. Adjusted datasets with a ratio within 30% of equality (0.7 - 1.3) were considered most plausible. This first validation allowed variations in the geographic distribution of malaria but set a reasonable reference point for the district average.

#### Distance Decay

To limit bias due to geographic access to health care, we assumed that there should not be an exponential distance decay relationship in adjusted malaria incidence (as it was observed in unadjusted incidence estimates, Figure S1). To assess this criterion, we calculated the ratio of average incidence in Fokontany located fewer than 5 km from a health center to average incidence in Fokontany more than 5 km away. Adjusted datasets with a ratio near 1 (0.7 - 1.3) were considered as most plausible. This second validation ensured that the geographic distribution of malaria incidence in the adjusted dataset was not associated with heterogeneities in geographic access to health care.

#### Financial Access

To limit bias due to financial access to health care, we assumed that average adjusted incidence in the catchment of health centers that implemented user-fee exemptions should be similar to those for which user fees were in place. Before adjustment, average monthly incidence of malaria among all individuals and children under five *inside* the PIVOT catchment area after financial barriers to care were removed were 13 and 27 per 1000 population, respectively, while the average monthly incidence among all individuals and children under five living *outside* of this area was significantly lower: 6 and 16, respectively (ratio of 2.1 and 1.7). To assess this criterion, we estimated the ratio of average adjusted malaria incidence in the catchment of health centers with user-fee exemptions to health centers without user-fee exemptions. Adjusted datasets with a ratio within 30% of equality (0.7 - 1.3) were considered as most plausible. This third validation ensured that the temporal and geographic distribution of malaria incidence in the adjusted dataset were not associated with heterogeneities in financial access to health care.

#### Geographic Heterogeneity

To assess the consistency of heterogeneities in malaria geographic distribution, we assumed that during the high transmission season (December to May) there is a geographic overlap with the distribution of reported fever in household surveys (April-May). In the high transmission season, 36.6% of individuals of all ages and 41.2% of children under 5 years presenting to health centers had a confirmed malaria diagnosis. Since malaria makes up a high proportion of febrile cases during these periods, geographic variations in febrile prevalence should be correlated with variations in malaria transmission. To assess this criterion, we estimated average fever prevalence for each of the 80 clusters in the I-HOPE cohort, and average malaria incidence for each of the 195 Fokontany during the high transmission season. Then, SaTScan software using the Bernoulli spatial model was used to identify geographic clusters of malaria in Ifanadiana district. SaTScan has been used in previous studies to identify spatiotemporal variation of malaria^45^ and other illnesses such as diarrheal disease^46^, schistosomiasis^47^, and colorectal cancer^48^. SaTScan cluster analysis was applied to identify spatial hotspots (i.e. higher than expected by random) among all individuals and among children under five in fever prevalence from survey data, unadjusted malaria incidence from health system data, and each of the adjusted incidence datasets. The area overlapped by geographic hotspots in fever and malaria from these different sources were quantified (Figure 3).

**Figure 3.**
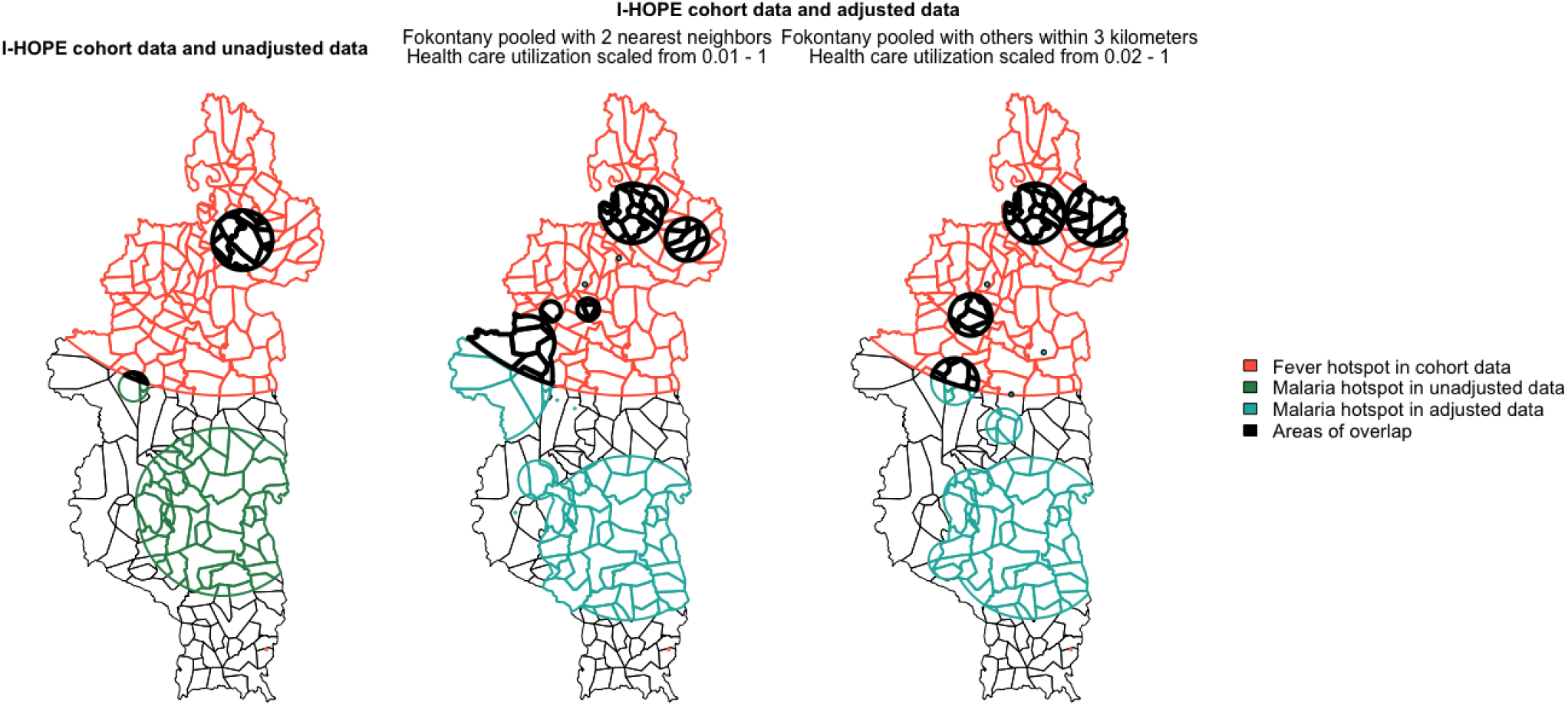
Comparison of geographic hotspots of fever (I-HOPE cohort survey) and malaria (health center registers) in Ifanadiana among all individuals during malaria high season, using unadjusted and adjusted estimates. Colored regions represent malaria hotspots in the various data sources and the areas of overlap in bold black. The left panel shows hotspots using unadjusted health center register data, while the center and right panels show examples of hotspots from two of the 168 adjusted datasets. The observed overlap is significantly greater in adjusted datasets, indicating improved consistency between the geographic distribution of fever in the I-HOPE cohort study and malaria in health center register data after adjustments.

All analyses were performed with R software, and R packages “lme4,” “gstat,” “rgdal,” “ggplot2,” “rsatscan,” “spdep,” “sp,” “rgeos,” “tidyr,” and “survey”^49^.

## RESULTS

### Malaria incidence in the unadjusted dataset and selection of the most plausible adjustment

Of the 314,443 patients who attended a health center in Ifanadiana district for an outpatient visit between 2014 and 2017, 270,747 patients had a known geographic location and came from within the district. Among these, 73,022 were confirmed malaria cases, 29,124 of which were children under 5 years. Average malaria incidence was 104.6 per 1000 population per year, and varied greatly across seasons. During the high transmission season (December to May), average malaria incidence was 168.0 per 1000 population per month, decreasing during the low transmission season to 41.3 per 1000 per month. There was a clear distance decay in malaria incidence both for individuals of all ages and for children under 5 years (Figure S1 in the appendix). Table 2 presents summary demographic and geographic characteristics of the patient population and malaria cases that attended one of the 19 health centers.

**Table 2.** Summary statistics of patient population in Ifanadiana health centers, 2014-2017.

|  | Population | Total patients | Total malaria confirmed cases | Malaria incidence per 1000 per year |
| --- | --- | --- | --- | --- |
| **Total** | 198,175 | 270,747 | 73,022 | 104.6 |
| **Age group** |  |  |  |  |
| Under 5 years old | 35,672 (0.18) | 92,533 (0.34) | 29,124 (0.40) | 231.8 |
| Over 5 years old | 162,504 (0.82) | 178,214 (0.66) | 43,898 (0.60) | 76.7 |
| **HSS catchment** |  |  |  |  |
| Inside | 72,152 (0.36) | 173,497 (0.64) | 42,992 (0.59) | 158.0 |
| Outside | 126,023 (0.64) | 97,250 (0.36) | 30,030 (0.41) | 70.5 |
| **Distance to health center** |  |  |  |  |
| 0-5 km | 63,811 (0.32) | 163,656 (0.60) | 41,067 (0.56) | 186.0 |
| 5-10 km | 81,787 (0.41) | 83,667 (0.31) | 25,294 (0.35) | 87.9 |
| 10-22 km | 52,577 (0.27) | 23,424 (0.09) | 6,661 (0.09) | 35.2 |

Of the 168 adjusted datasets evaluated for individuals of all ages (Figure 4), only one dataset fulfilled the four criteria described above (Table 3) and 86 datasets fulfilled three of the four criteria. Every pooling group and lower limit of utilization index was represented among the datasets that fulfilled three but not four criteria. We observed a clear trade-off in the adjusted datasets for the different evaluation criteria. Setting the lower limit for the utilization index at lower values (e.g. 0.01-0.07) resulted in better corrections for financial and geographic trends but overall incidence was above acceptable thresholds (Figure 4, Figure S7). In contrast, setting the lower limit for the utilization index at higher values (e.g. ≥ 0. 09) resulted in overall incidence closest to incidence in the Fokontany with optimal access to care, but there remained important bias due to financial and geographic access (Figure 4). The most plausible dataset was obtained using a lower limit of 0.08 for the health care utilization index in the benchmark multiplier method, and pooling Fokontany with two nearest neighbors. Figure 5 shows how the adjustment in this dataset improved geographic and temporal patterns in malaria incidence, reducing the apparent difference between Fokontany inside and outside of PIVOT intervention following user-fee removal, and removing the distance decay observed in the unadjusted dataset.

**Figure 4.**
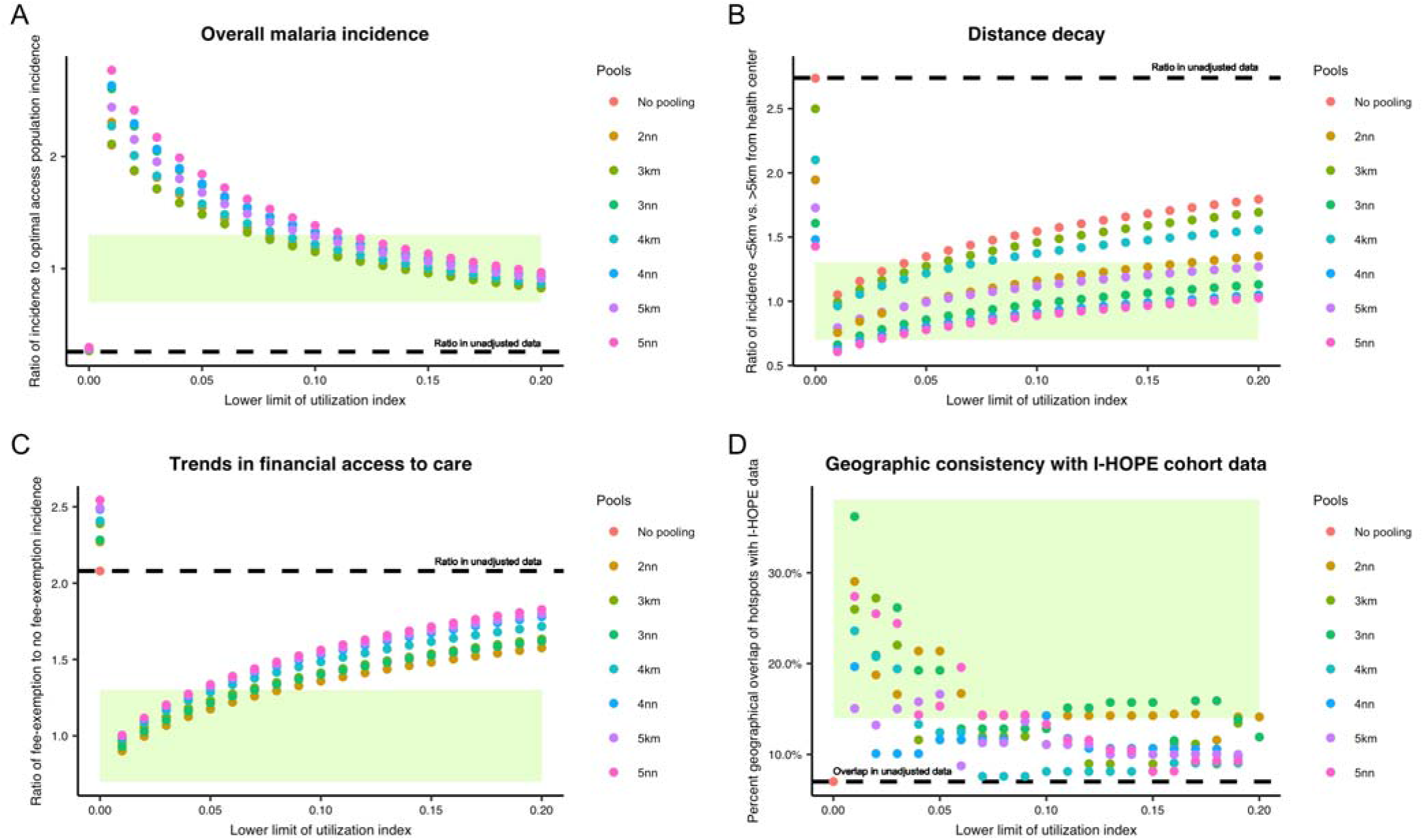
Summary results for the four evaluation criteria in unadjusted data and all adjusted malaria datasets. Each dot represents the metric of interest in one set of adjusted data, and colors represent the pooling strategy (e.g. 2nn = pooling of Fokontany with its 2 nearest neighbors; 3km = pooling with neighbors within 3km). The dashed line shows values for the unadjusted dataset. Shaded green areas show target ranges of each evaluation criteria as described in Table 1. (A) Overall malaria incidence: ratio of malaria in adjusted dataset to malaria in optimal access areas. Values closer to 1 mean better performance. (B) Distance decay: ratio of incidence in Fokontany less than 5 km from a health center to incidence in Fokontany more than 5 km from a health center. Values closer to 1 mean better performance. (C) Trends in financial access to care: ratio of average monthly incidence in fee-exempt to non-fee-exempt populations in each adjusted dataset. Values closer to 1 mean better performance. (D) Geographic consistency with I-HOPE cohort data: percent of overlap between hotspots of fever identified in the I-HOPE cohort study data and malaria incidence in each adjusted dataset. Greater values mean better performance. Equivalent plots including analyses for children under 5 years can be found in the appendix.

**Table 3.** Summary results for the four evaluation criteria in unadjusted data and best-performing adjusted malaria dataset for individuals of all ages. An equivalent table for children under 5 years can be found in the Appendix.

| Dataset | Ratio of average incidence in dataset to incidence in optimal access areas | Ratio of incidence < 5km to > 5km from a health center | Ratio of incidence in fee-exemption to non-fee-exemption areas in dataset | % of hotspot clusters overlapped between dataset and I-HOPE cohort data |
| --- | --- | --- | --- | --- |
| Unadjusted register data | 0.26 | 2.74 | 2.08 | 7% |
| 2 nearest neighbors, utilization index 0.08 – 1 | 1.29 | 1.10 | 1.29 | 14% |

**Figure 5.**
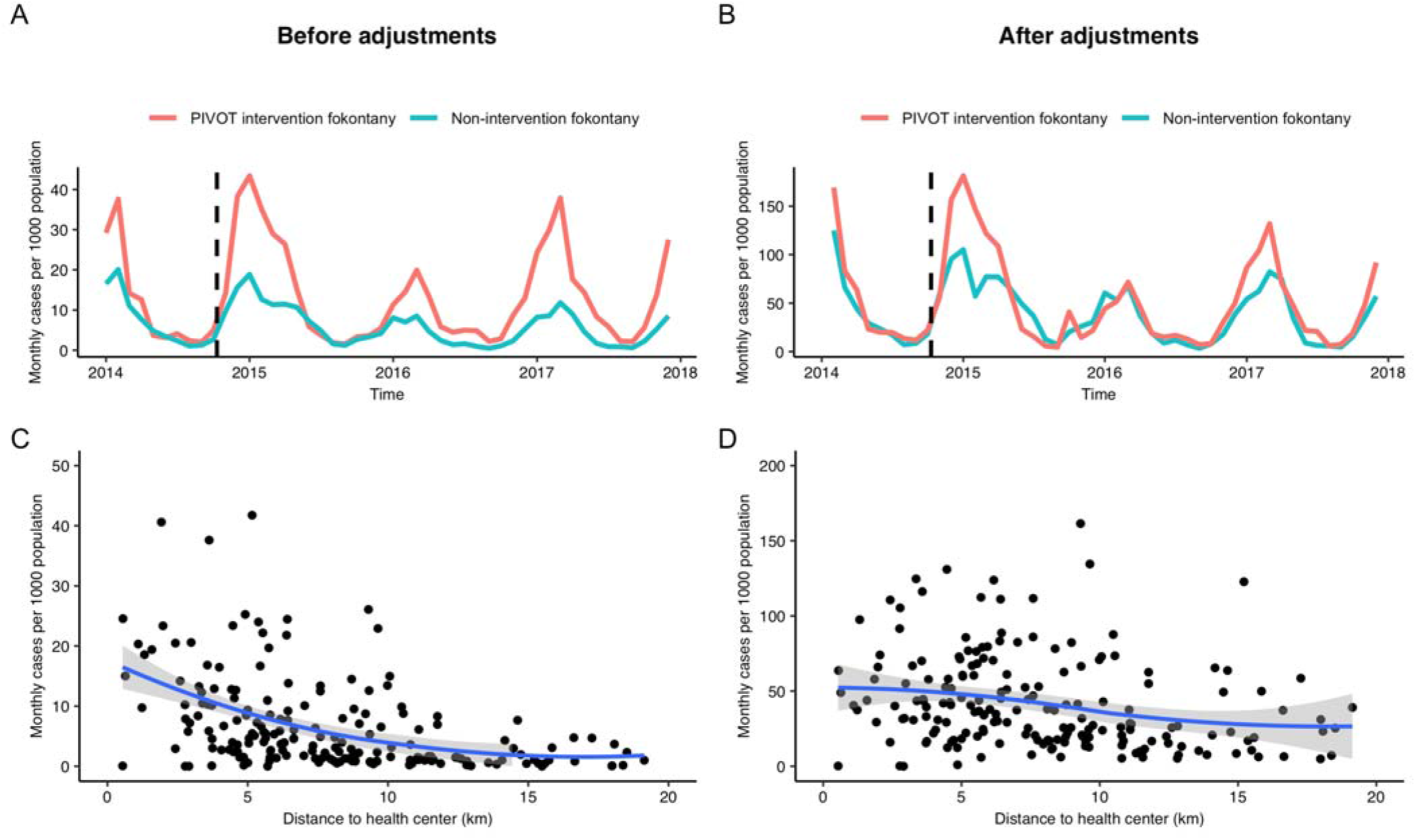
Temporal and geographic patterns in malaria, before and after adjustments. The top two panels show the average monthly cases per 1000 population over time, with colors representing the PIVOT intervention (orange) and nonintervention (teal) Fokontany, (A) before and (B) after adjustments in the most plausible dataset. The vertical dashed lines indicate the date (October 2014) when user fees were removed from health centers in PIVOT intervention Fokontany. The bottom two panels show the average monthly malaria cases per 1000 population in each Fokontany by distance to the nearest health center, (C) before and (D) after adjustments for health care access. Solid lines are the smoothed conditional means (LOESS method) and grey areas are the 95% confidence interval around the mean. Equivalent plots including only children under 5 years can be found in the appendix.

For children under five, 13 datasets satisfied the four criteria. The lower limits of utilization were higher than for individuals of all ages, ranging from 0.14 to 0.2 (Table S3, Figure S4). Similar to the trends among all individuals, setting the lower limit of the utilization index at lower values (0 - 0.15) improved corrections for financial and geographic trends, but resulted in unacceptably high overall incidence. Datasets with high utilization index values (0.15 - 0.2) and low pooling groups (2-3 nearest neighbors) performed best overall. The most plausible dataset was obtained using a health care utilization index rescaled from 0.19 to 1 in the benchmark multiplier method, and pooling Fokontany with three nearest neighbors.

### The hidden burden of malaria in Ifanadiana

Using adjusted incidence estimates from the most plausible dataset, we estimated that the number of malaria cases diagnosed via passive surveillance in Ifanadiana from January 2014 to December 2017 represented only 21% of the total number of cases that could have occurred among all individuals during the study period, and 32% among children under 5 (Table 4). Average adjusted malaria incidence was estimated at 518 per 1000 population per year for individuals of all ages (43 per 1000 per month) and 733 per 1000 population per year for children under 5 (61 per 1000 per month). Average adjusted malaria incidence per month was nearly four times higher during the high transmission season (70 per 1000) than during the low transmission season (18 per 1000).

**Table 4.**
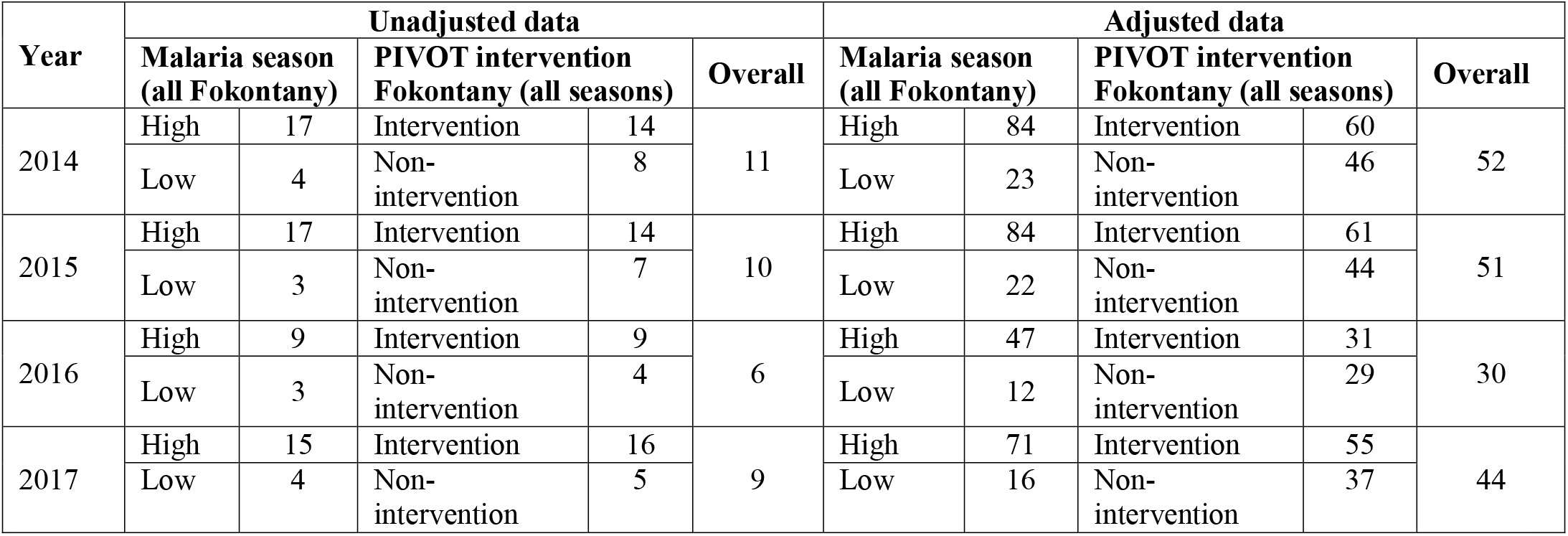
Average monthly incidence of malaria for individuals of all ages in Ifanadiana from unadjusted and adjusted data, by transmission season and PIVOT intervention area, in cases per 1000 population. An equivalent table for children under 5 years can be found in the Appendix.

Temporal dynamics in the adjusted dataset showed a decrease in malaria incidence from 2014-2015 (613 cases per 1000 per year) to 2016-2017 (441 cases per 1000 per year), with peaks in monthly incidence decreasing from almost 150 to about 100 cases per 1000 respectively (Figure 6A). This trend is observed to a lesser degree in the unadjusted data, but when unadjusted data is disaggregated by intervention area, incidence in PIVOT intervention areas appear to have increased since 2014, likely due to increased access to care in these areas. After adjustments, the average monthly incidence of malaria is higher overall and more stable over time and between intervention and non-intervention areas due to adjustments for changing health care utilization (Figure 6A).

**Figure 6:**
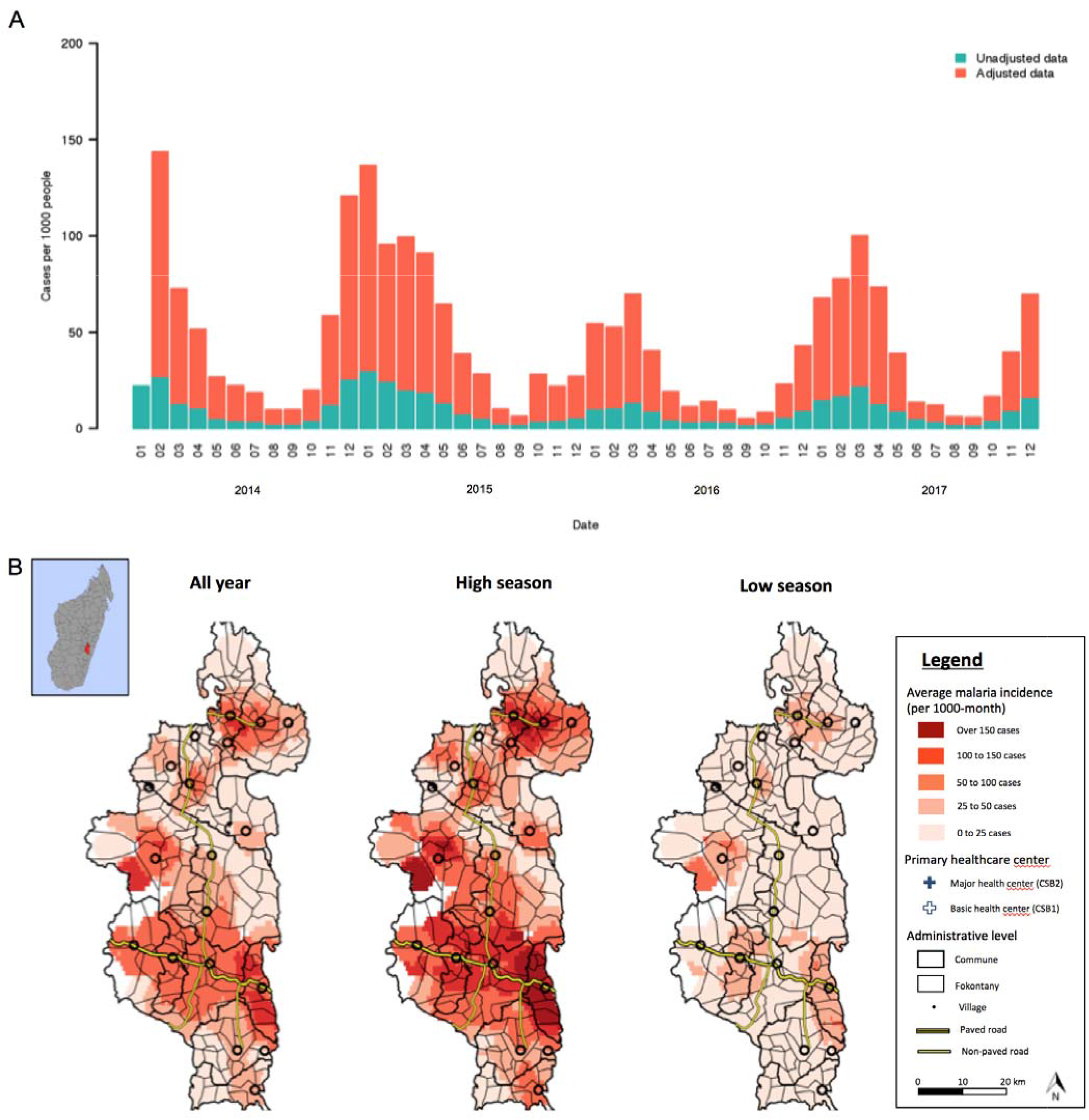
Temporal and spatial dynamics of adjusted monthly malaria incidence in Ifanadiana, 2014-2017. (A) Average number of new cases per 1000 population of all ages per month in the most plausible adjusted dataset (orange) and before adjustment (teal). An equivalent plot including only children under five can be found in the appendix. (B) Geographic distribution of malaria, averaged over all months (left), high season months (December to May; center), and low season months (June to November; right). Color gradient represents average monthly malaria incidence per 1000 population. Equivalent plots of spatial distribution in unadjusted health center register data are included in the appendix.

Geographic distribution in the adjusted dataset revealed clusters of high incidence in low elevation areas in the northeast and southeast of the district (Figure 6B). In addition, another high incidence cluster was observed in the western part of the district, at higher elevation and close to the limits of Ranomafana National Park. These high transmission clusters were stable across transmission seasons (Figure 6B) and years (Figure S2). In addition, 5% of Fokontany in Ifanadiana district had an average incidence higher than 100 cases per 1000 per month, distributed mostly in the central and southern part of the district (Figure 6B). In comparison, the unadjusted dataset only revealed areas of high incidence in very close proximity to health centers along the main paved road and with user-fee exemptions in place (Figure S3), missing most relevant transmission areas. Detailed spatio-temporal dynamics of malaria per month, from both the unadjusted and the most plausible adjusted dataset can be visualized in Video S1.

## DISCUSSION

Despite the increasing use of disease modeling and precision health mapping to inform national or regional health planning, their application remains scarce at the local level, where intervention efforts actually take place. This is especially true in rural areas of sub-Saharan Africa where the burden of infectious diseases is the highest. Improving the quality of routine surveillance data is critical for identifying at-risk populations and targeting resources in order to achieve universal access to diagnostics and treatment, which could contribute to the elimination of endemic diseases like malaria^50^. Here, we propose a method to improve existing passive surveillance data using models of health care utilization in order to produce more realistic estimates of local disease incidence over space and time. Using the example of malaria in a poor rural district of Madagascar, we show that adjusted incidence estimates were less biased by differences in financial and geographic access to health care between populations. We estimated that passive surveillance in Ifanadiana could have missed about 4 in every 5 cases of malaria individuals and 2 out of every 3 cases among children under five.

Passive surveillance systems are a cornerstone of many disease control programs because they are relatively inexpensive and can efficiently cover large geographic areas. When access to health care is relatively homogenous in a country, variations in incidence across districts help control programs identify those with higher transmission and allocate resources accordingly^51,52^. However, at the local level of a health district these systems are systematically biased towards areas of good health care access (e.g. near health centers), preventing the implementation of geographically targeted interventions in areas of high transmission. Active surveillance systems, on the other hand, can capture a significantly higher proportion of cases and produce more accurate incidence estimates. Unfortunately, in the case of malaria they are too expensive to be used routinely in areas of high transmission, and the results cannot be extrapolated to detect variations in malaria in regions outside of the study area or period^13–18^. Thus, our study fills a significant gap for malaria surveillance, which could be applicable to other diseases. Using existing passive surveillance data, we were able to produce spatially-explicit estimates of malaria incidence for every community within a health district over time, identifying hotspots of transmission in communities with poor health care access that were previously invisible from passive surveillance. This could help inform local program implementation in Ifanadiana and similar high transmission settings without requiring extensive resources.

Without improvements to passive surveillance strategies, countless preventable cases and deaths of malaria may continue to take place and go unnoticed, which could undermine goals set for a 90% reduction in malaria mortality and the elimination in at least 35 countries by the year 2030^12^. Our results suggest that only 21% of malaria cases were detected by routine passive surveillance in Ifanadiana district. This is consistent with findings from other settings where active and passive malaria surveillance methods were compared. For example, a study in rural Kenya found that the incidence of malaria in children was over three times higher when active surveillance was used compared to passive surveillance^15^. A similar study in central India reported that malaria incidence was almost eight times higher when calculated using active rather than passive surveillance data^16^. In 2012, the World Health Organization estimated that only 14% of malaria cases globally were captured by routine surveillance^17^. In Ifanadiana, a mountainous landscape, poor road infrastructure and a sparsely distributed population make it difficult for patients to access health centers. More than 95% of paths are not accessible by vehicle, and three fourths of the population live more than an hour’s walk of a public health center^36,37^, a commonly accepted threshold of low geographic access^53-56^. All these factors could have led to significant underreporting of malaria, at levels compatible with estimates presented here.

After adjustment, we observed significant spatial variations in malaria incidence in communities across the district, with 7% the population (13,201 people) living in areas where annual incidence was twice the district’s average. Although malaria heterogeneity and its drivers are commonly modelled at the national and regional level^57-59^, malaria can have extensive spatial variability in relatively small areas^43,44,60^. For instance, fine-scale variations in socio-demographic and behavioral factors can influence malaria risk in remote communities^61^ or affect adherence to malaria control programs^62^. Moreover, local variations in environmental factors such as temperature, rainfall, land cover, and altitude have been shown to influence malaria geographic distribution^63-65^. Fine-scale estimates of malaria spatio-temporal variations obtained here can then be used to characterize local socio-economic and environmental drivers of malaria risk, paving the way to the development of forecasting systems that could guide local malaria control.

Although our study was retrospective and we had to collect information directly from paper registers, which was extremely time and resource consuming, this approach could be scaled-up in the future to other settings and diseases that rely on passive surveillance. Indeed, a push for electronic data collection to improve health information systems is underway at health care facilities of developing countries, with the current scale-up of the open source DHIS2 (District Health Information Software)^66^ among other platforms. These platforms can be combined with mobile tools for registering cases and track patient-level data at different levels of care. The level of granularity and timeliness of data that these e-health platforms offer when compared with traditional health management and information systems (e.g. paper-based registries, monthly aggregation in electronic databases) opens new possibilities for disease control which are still largely unexplored. In particular, integration of feedback loops between disease modelling approaches and e-health surveillance platforms could help to 1) target efforts and plan resources necessary ahead of time for specific areas and periods, reducing stock-outs and increasing case detection; and 2) implement additional control activities that are predicted to minimize transmission at the population level.

This study had several limitations. First, there was no active surveillance campaign during the study period that could serve as a true comparison point for selecting the most plausible set of estimates. As an alternative, we compared adjusted estimates with areas within the district that had optimal access to care and therefore were assumed to have missed few malaria cases. However, if these areas were not representative of overall malaria incidence due to heterogeneities, this could have resulted in an under- or overestimation. Second, many of the most remote Fokontany did not report any malaria cases even during high transmission seasons. To allow for adjustments and minimize underestimation of malaria in these remote populations, we pooled these Fokontany with their nearest neighbors, but this likely reduced the spatial precision of our estimates. Third, even though we correct for health care access, there were still some patterns in the adjusted datasets (e.g. higher incidence around PIVOT-supported health centers), which could suggest an influence of unmeasured factors not accounted for in our analyses. Finally, although data on RDT stock-outs was available, underreporting of the number of days without stocks in some health centers could have led to artificially low malaria estimates. Despite its limitations, we are not aware of any other study that has attempted to systematically address sources of malaria underreporting to generate realistic incidence estimates from local passive surveillance systems.

In conclusion, although passive surveillance at health facilities remains the prevailing surveillance system for many endemic diseases in the developing world, systematic biases in these data prevent their use to inform local disease control programs within health districts. By adjusting for health care access and other sources of underreporting, we show that passive surveillance can be used to obtain realistic estimates of malaria dynamics with a level of spatial resolution that is locally actionable. Future research should assess whether such methods can be scaled-up and integrated with e-health platforms currently being deployed.

## Data Availability

Data can be made available upon request at

## ACKNOWLEDGEMENTS

We are grateful to everyone who contributed to the participatory mapping of Ifanadiana, especially Vincent Herbreteau, Christophe Révillion, Jérémy Commins, and Blake Girardot. We thank the staff of the local Ministry of Health team in Ifanadiana district as well as PIVOT’s monitoring and community teams for their support during data collection. Thanks are due to Benjamin Andriamihaja, Benjamin Roche, and Mauricianot Randriamihaja for their help at different stages of the project.

## AUTHOR CONTRIBUTIONS

Conceived and designed the experiments: EH, MHB, AG. Performed the analysis: EH, FAI, AG. Contributed reagents/materials/data/analysis tools: ACM, MR, FN, MAN. Wrote the initial draft of the manuscript: EH, AG. Revised the manuscript and accepted it in its final form: EH, MHB, FEI, ACM, LFC, MR, MB, FN, MAN, AG.

## SUPPLEMENTARY INFORMATION

**S1 Supplementary information**. It contains 6 supplementary figures and 2 supplementary tables, with results for children under 5 years and other additional information.

**S2 Video of malaria spatio-temporal dynamics**. It shows geographic changes in monthly malaria incidence in Ifanadiana district (unadjusted and adjusted estimates from the most plausible dataset). For reference, removal of user fees in the initial HSS intervention catchment (in red) took place in October 2014, and was expanded to one additional commune in October 2017.

